# Under Pressure: The Burden of Hypertension on Armenia’s Ambulance System

**DOI:** 10.64898/2026.07.22.26358725

**Authors:** Ani Arzoumanian, Anya Agopian, Marine Hovhannisyan, Aline Baghdassarian, Sharon Chekijian

## Abstract

Hypertension is a leading cause of premature mortality globally. Armenia, a country located in the South Caucasus, has one of the highest rates of hypertension in the world, with an estimated prevalence of 51% among adults aged 30-79. A previous analysis of ambulance calls from 2016-2022 revealed that high blood pressure was the most common ambulance complaint in Armenia. In the present study, we analyzed a de-identified database of all ambulance calls logged on the Locator™ software in Armenia from January 1, 2016 to July 31, 2022. Among all adults who reported at least one chief complaint, 28.8% listed high blood pressure as their sole complaint. Within this cohort, 61.0% of patients were over 60 years old, and 70.1% of patients were women. The rate of transfer to the hospital was 9.2%. Call volume fluctuated by season, with higher frequencies in the winter and spring and lower frequencies in the summer and fall. Results indicate that a significant proportion of ambulance calls are for non-emergency cases of high blood pressure with no additional reported medical complaints, indicating that Armenia’s elevated prevalence of hypertension places a significant burden on its ambulance system. Policymakers should prioritize hypertension as a national issue, and consider multi-modal policy interventions aimed at reducing ambulance utilization for high blood pressure.

## Background

Noncommunicable diseases (NCDs) account for over half of the global burden of disease. Cardiovascular disease (CVD) in particular has been identified as a leading cause of death, accounting for half of all mortalities attributed to NCDs.(1,2) The global prevalence of hypertension, which is the largest single risk factor for CVD mortality and causes an estimated 11 million avoidable deaths annually, is on the rise, and today affects 34% of adults aged 30- 79.(3,4) In the resource-constrained environments of low- and middle- income countries (LMICs), NCDs constitute a significant macroeconomic strain, draining national resources that could otherwise be invested in health system strengthening.(5–7)

The Republic of Armenia is a land-locked country located in the South Caucasus. Over the last 30 years since independence, Armenia has progressed from low income to upper-middle income country classification.(8) In the face of this rapid evolution, Armenia continues to face many challenges with modernizing its healthcare systems, including mitigating growing NCD rates among its population. The World Health Organization estimates that 21% of Armenia’s population are at risk of premature death due to NCDs, which also constitute an economic burden equivalent to 6.5% of Armenia’s GDP.(4,9) An estimated 51% of adults aged 30-79 in Armenia have hypertension. Among them, only 44% are diagnosed, 32% are treated, and 10% achieve blood pressure control. These findings are supported by yearly hypertension opportunistic screening studies in Armenia.(10–12) Thus, Armenia faces a much higher prevalence of hypertension than most countries in the world, regardless of income level or region, including all four of its neighboring countries (Turkey, Iran, Georgia and Azerbaijan).

In accordance with recent World Health Organization directives to integrate emergency, critical and operative care, Armenia’s Emergency Medical Services (EMS) system has been identified as an area needing significant improvement to advance overall national health metrics.(13–17) Armenia’s EMS system follows the Franco-German (stay and stabilize) model, in which the majority of emergency care is delivered by ambulance doctors and nurses out-of-hospital. In Armenia, ambulances are fully subsidized by the government, making them free of charge for all patients. Call data are input into Locator™, a software implemented in all 54 state-run ambulance stations in 2011.

An analysis of ambulance calls from 2016-2022 by this team revealed that high blood pressure was listed as a complaint in 26.4% of calls, making it the most common ambulance complaint in both the capital city (Yerevan) and rural provinces (Marzes).(13) This finding prompts a need for further investigation of cases of high blood pressure within the EMS system. In this retrospective analysis, we evaluate and describe ambulance calls related to high blood pressure among adults in Armenia and utilize a systems-lens to provide policy recommendations directed toward mitigating the effects of disproportionate hypertension calls on the EMS system in Armenia.

## Methods

De-identified data from Locator™ was provided by the Ministry of Health of Armenia. Data included all ambulance call entries from January 1, 2016 to July 31, 2022. All data was downloaded on July 31, 2022. Descriptive statistics were analyzed using IBM SPSS Statistics, version 28.0.1.1 (14). Non-numerical values in the database were provided in the Armenian language, which the authors of this paper read and translated. Analyses of demographic information, such as age, gender, or transport to hospital, included only those calls which reported this information. Chief complaints are reported by the caller or patient, and given as the reason for requesting an ambulance. Categorial variables are presented with frequencies and proportions.

Fourteen of Armenia’s 54 ambulance stations were excluded from consideration due to missing data (see Arzoumanian et al., 2024 for more information).(13) Nonsensical outliers in age and date columns were removed and counted as missing information. Age was limited to the range of 0-100 years old. Analysis was limited to patients 18 years and older. Seasons were classified as Winter (December-February), Spring (March-May), Summer (June-August), and Fall (September-November).

## Results

### Patient Demographics

Adults (age 18+) constituted 92% (n=2,022,218) of all ambulance patients. Among adults, patients over 60 years old represented the largest age demographic (49.2%), followed by patients aged 40-59 (29.6%) and 18-39 (21.1%).

Among all adults who reported at least one chief complaint, 28.8% (n=406,152) listed high blood pressure as their sole complaint, while an additional 0.3% of callers listed high blood pressure alongside one or more additional complaints. Among those who listed high blood pressure as their sole complaint, 61.0% of patients were over 60 years old, 32.3% were aged 40- 59, and 6.8% were 18-39 (Table 1). A majority of both male (56.6%) and female (63.3%) callers were over 60 years old. Women accounted for 70.1% of calls in which gender was listed. Women over 60 years old accounted for 44.4% of calls, followed by women aged 40-59 (22.3%) and men over 60 years old (17.0%).

**Table 1.** Age and Gender Demographics of Patients with High Blood Pressure as Sole Complaint.

|  |  |  | Gender |  |  | Total |
| --- | --- | --- | --- | --- | --- | --- |
|  |  |  | Male | Female | Gender Not Listed |  |
| Age | 18-39 | Freq. | 7,643 | 9,739 | 10,435 | 27,817 |
|  |  | % within gender | 9.1% | 5.0% | 6.8% | 6.8% |
|  | 40-59 | Freq. | 28,806 | 62,460 | 39,341 | 130,607 |
|  |  | % within gender | 34.3% | 31.8% | 32.2% | 32.2% |
|  | 60+ | Freq. | 47,460 | 124,450 | 75,818 | 247,728 |
|  |  | % within gender | 56.6% | 63.3% | 60.4% | 61.0% |
| Total |  | Freq. | 83,909 | 196,649 | 125,594 | 406,152 |
|  |  | % within gender | 100.0% | 100.0% | 100.0% | 100.0% |

### Additional Findings

Among adult patients who listed high blood pressure as their sole complaint, the rate of transfer to the hospital was 9.2%.

Call frequency was assessed by location over time. There was a substantial drop in overall ambulance call reporting in the database from Yerevan in 2017-2019, with many months listing 0 high blood pressure calls (Figure 1).

**Fig 1.**
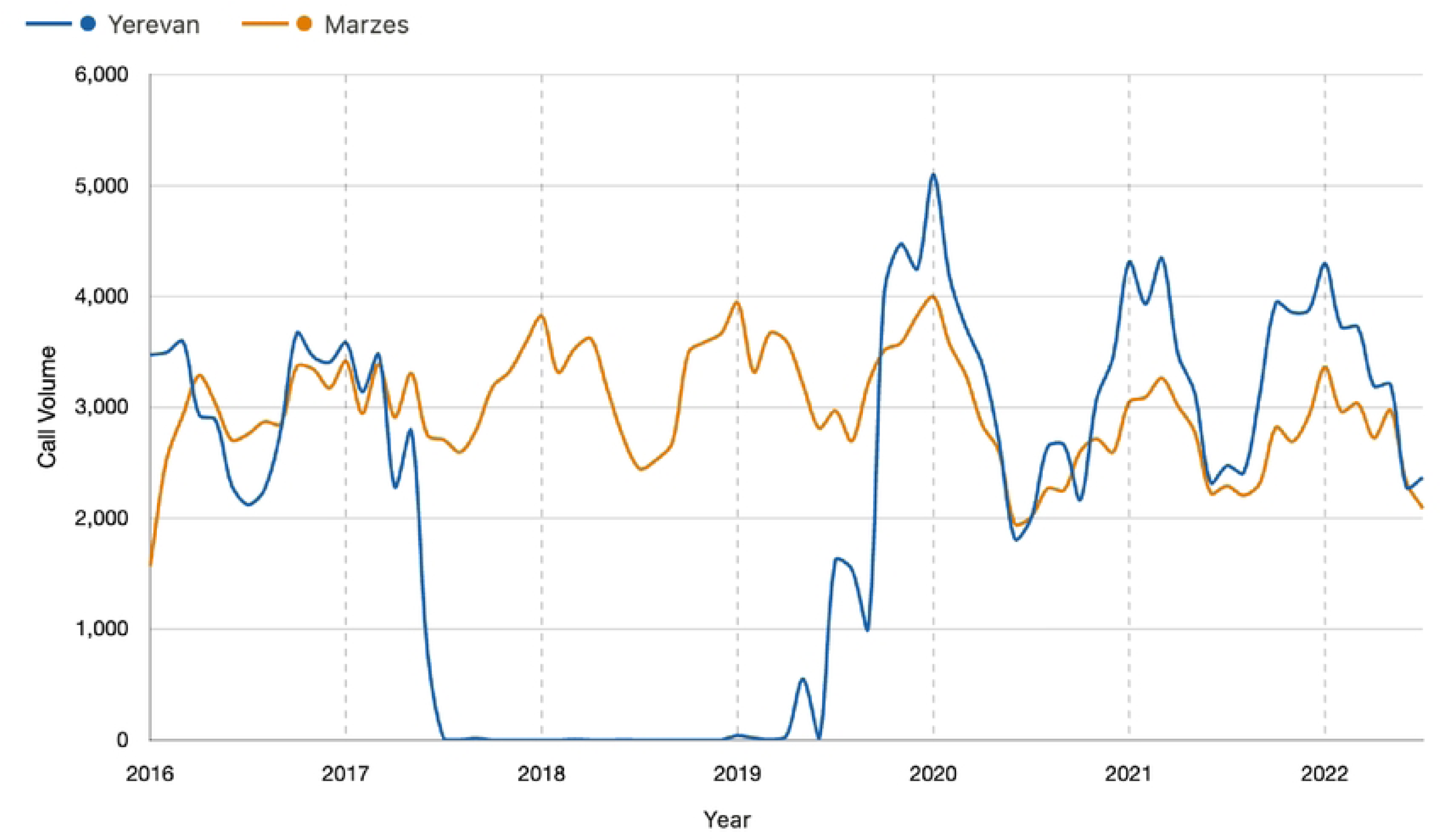
High Blood Pressure Calls Over Time by Location.

Call volume significantly fluctuated by season (Figure 2). Volume was higher in the winter (29.3%) and spring (28.5%) compared to summer (19.4%) and fall (22.8%). This pattern was generally consistent across all years and regions; however, seasonal changes could not be assessed in Yerevan alone due to the missing data described above in 2017-2019.

**Fig 2.**
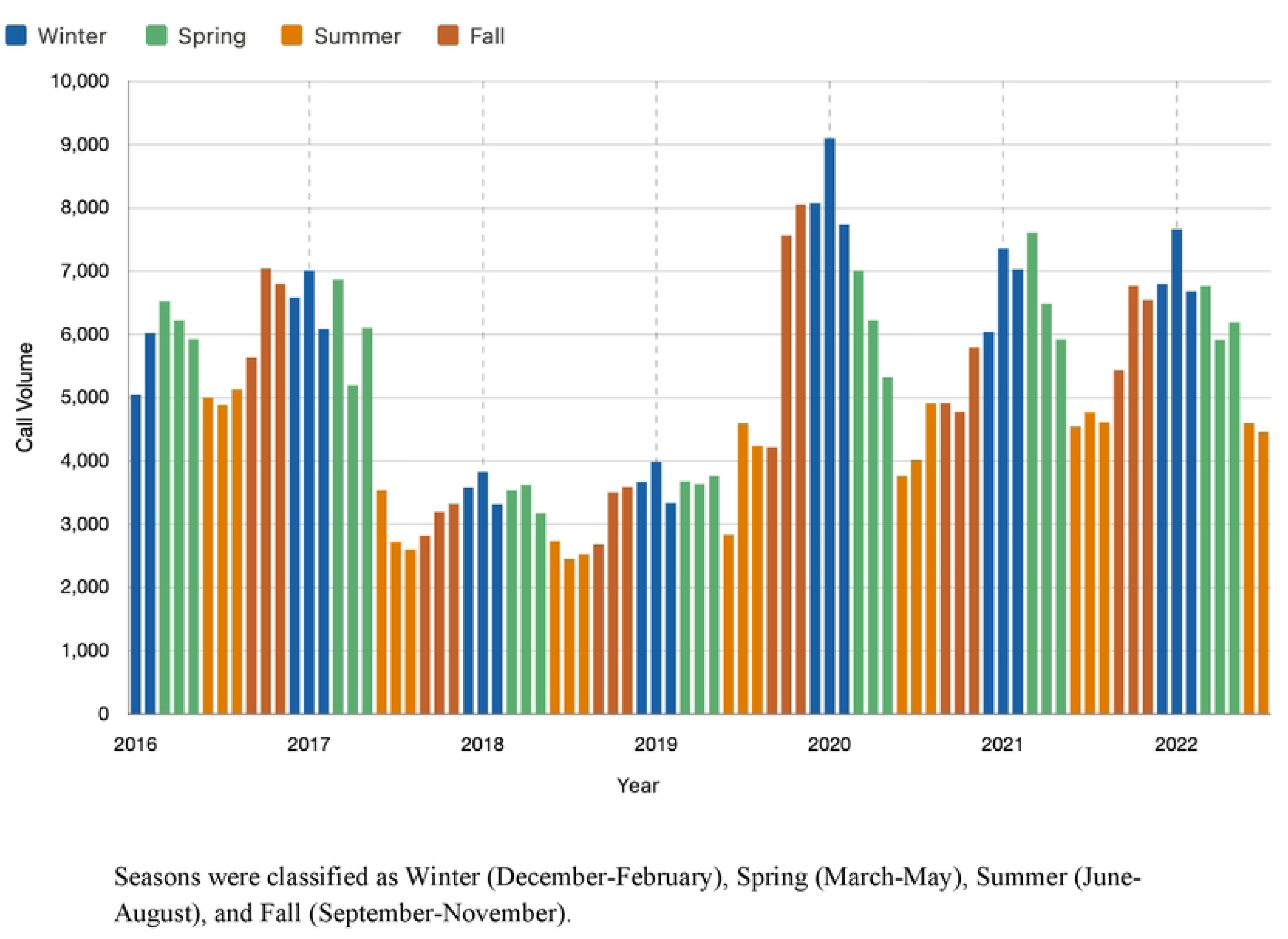
High Blood Pressure Calls by Season.

## Discussion

### Armenia’s Hypertension Burden

The overall burden of hypertension in Armenia’s population is well documented; however, the impacts on health systems and EMS capacity have not been adequately studied. In the present study, we find that nearly 29% of adult ambulance callers listed high blood pressure as the sole reason for calling an ambulance, contributing a remarkable burden on Armenia’s ambulance system. The frequency of these calls increases steeply with age. To our knowledge, no other country has reported high blood pressure as a common ambulance complaint, indicating that this finding is unprecedented by global standards. Moreover, we find that only 9.2% of cases result in ambulance transport to a hospital, a much lower rate than the previously reported 17.6% rate of transfer for all ambulance calls in Armenia.(13) The low transport rate, paired with the finding that only 0.3% of high blood pressure callers listed additional complaints, implies that the vast majority of patients calling for high blood pressure likely do not meet criteria for further evaluation and thus are not truly emergency calls as they would be defined by a physician.

Ambulance services are free to the public, with costs covered by Armenia’s Ministry of Health (MOH) even prior to the passing of universal health coverage (UHC) on January 1, 2026. The low transport rate found in the present study implies that Armenia’s ambulance system is likely heavily utilized for non-emergency complaints, raising the question of moral hazard.

Authors of the present study conducted extensive ambulance ride-alongs in Armenia to understand processes related to EMS calls as background for this study. Anecdotally, the culture in Armenia normalizes the possession and frequent usage of blood pressure cuffs in the home.

Though this practice is not well documented in the literature, it is a common observation in Armenia and may help to contextualize the findings of the present study. Many patients calling an ambulance for blood pressure likely do so after measuring elevated blood pressures themselves at home. Importantly, these measurement devices are likely not recalibrated, and patients are not commonly trained to measure or interpret blood pressure.

These results and observations raise concern that EMS is being utilized in place of primary or urgent care visits. For patients who do utilize EMS as their primary support for managing high blood pressure, ambulance intervention (via single dose of blood pressure medication) would be largely ineffective, and may even contribute to poorer health outcomes, as studies have shown that blood pressure variability is associated with increased risk of cardiovascular diseases, stroke, and all cause mortality. (18)

### Gender Demographics

Data from the WHO indicate that men in Armenia have increased risk factors for developing hypertension due to salt intake, physical inactivity, and tobacco and alcohol use compared to women.(2,4) Armenian men have a similar hypertension prevalence (51%) as Armenian women (50%); yet, men are less frequently diagnosed (39% vs 49% for women), less frequently treated (25% vs 37% for women), and less likely to achieve blood pressure control (9% vs 11% for women). Interestingly, these findings stand in stark contrast with the results of the present study, which reveal that women are 2.3 times more likely to call an ambulance for blood pressure than men, despite having increased diagnosis, treatment, and control of hypertension overall. This phenomenon may be explained by more treatment-seeking behavior by women, in both a primary care and emergency setting, compared to men.

### Age Demographics

Data on age demographics for patients with hypertension in Armenia is limited; however, the finding in the present study of age-related increase in frequency of high blood pressure calls is expected given increased prevalence of hypertension in older populations. It is likely that additional factors, including comorbid medical conditions, decreased mobility, decreased access to transport, and financial barriers, play a role. In the present study, patients over age 60 accounted for 61% of ambulance calls for high blood pressure, indicating that this age demographic in particular should be targeted for intervention (e.g. screening, education, and removal of financial barriers to outpatient care).

In January 2026, Armenia rolled out the first phase of its universal health coverage (UHC) plan. Notably, Phase 1 of the UHC covers vulnerable populations (encompassing approximately 1.6 million people), including senior citizens aged 65 and older.(19) Health policy interventions targeting this demographic, in particular, have the potential to significantly reduce the burden of high blood pressure calls on ambulance systems.

### Seasonal Fluctuations

Frequency of high blood pressure calls fluctuated by season. Frequency was higher in the winter and spring compared to the summer and fall. Winter months had the highest burden, with 29.3% of high blood pressure calls occurring in that season, while summer had the lowest frequency, with 19.4% of calls occurring in those months. This observed difference between seasons is expected, and likely has multiple causes.

Blood pressure commonly exhibits variability due to changes in physical activity, sleep, and temperature.(20) Seasonal fluctuations have been previously documented — notably, blood pressure tends to increase in the winter, which has largely been attributed to cold weather, and decrease in the summer. Additionally, health risk factors such as salt intake, tobacco and alcohol use, and physical inactivity may also fluctuate by season; however, these patterns have not been documented in the literature for Armenia.

### Policy Recommendations

The burden of hypertension on Armenia’s ambulance system is significant and seemingly unparalleled in the global literature. Policy reform should focus on both addressing the overall prevalence of hypertension in the population, in accordance with global recommendations, and also specifically reducing the burden of hypertension calls on EMS.

### Hypertension reduction policies

The WHO, in its Global Report on Hypertension (2025), outlines five recommendations for rapid and sustainable prevention, treatment, and control of hypertension, with the overall aim of reducing cases of stroke, myocardial infarction, kidney disease, and dementia, as well as the associated socioeconomic burdens of these diseases.(4) The guidelines direct countries to: 1) Integrate hypertension interventions into universal health coverage, 2) Improve access to affordable antihypertensive medications and validated measurement devices, 3) Invest in the health workforce and expand team-based care, 4) Monitor population trends via strengthening health information systems, and 5) Raise public awareness of the silent dangers of hypertension.

Armenia’s recent adoption of its UHC plan makes it an opportune moment to incorporate robust hypertension coverage policies. These would include subsidization of hypertension screening, primary care visits, and antihypertensive medications. Moreover, Armenia’s MOH must ensure the availability (both financial and physical accessibility) of a variety of first and second line antihypertensives, which are vital for the achievement of adequate pressure control.

The MOH should additionally consider a stronger approach in addressing risk factors to reduce the overall prevalence of hypertension. These include salt intake, tobacco and alcohol use, obesity, and physical inactivity. Although these interventions are unlikely to reduce the burden on EMS in the near term, they are part of a comprehensive strategy to address population level concerns long term.

### Reducing the burden of high blood pressure calls on EMS

In the context of Armenia’s ambulance system, the present study provides evidence of widespread public misuse of ambulance resources. This is a drain on government resources, which could otherwise be utilized for preventive interventions, screening, and treatment of hypertension. Moreover, non-emergency calls for high blood pressure reduce ambulance availability for those patients who do require acute treatment and transport to a hospital, potentially contributing to delayed interventions for time-sensitive conditions like stroke and myocardial infarction.

When specifically addressing the undue burden of blood pressure calls on EMS, policies aimed at reducing moral hazard could be highly effective. Policies to consider might aim at removing EMS coverage for calls for isolated complaints of high blood pressure without secondary complaints concerning for concomitant end organ threats (such as chest pain, stroke symptoms, headache, or shortness of breath). Such restrictions must be applied carefully to avoid the opposite effect of discouraging patients from seeking care for true emergency conditions and placing the burden of medical triage on the patient. Suitable triage tools at the time of the EMS call-in may play an important role here in discouraging undue burden on the EMS system. In this case, staff taking the initial call could be prompted to use validated screening questions to determine the proper course for the patient and whether or not to dispatch an ambulance versus referring the patient to their primary doctor or polyclinic. Given the sheer frequency of ambulance calls for high blood pressure, the MOH could pilot a hotline specifically for hypertension. Guidelines for appropriate dispatch of ambulance services and referral to primary care assessment and follow-up would support accurate clinical decision making and should greatly decrease this burden of hypertension on EMS.

Risk reduction, as outlined above, and patient education are paramount to overall efforts to reduce hypertension prevalence and the burden of high blood pressure calls on Armenia’s ambulance system. The MOH may consider utilizing a nationwide television broadcast to educate the population about high blood pressure and encourage patients to pursue diagnosis and treatment in a primary care setting. An alternative educational tool could be the creation of pamphlets for use by ambulance staff and by primary care doctors. These can be designed to be both informational (e.g. common causes, preventive strategies, and treatment options) and instructional (e.g. red flag symptoms and when to call an ambulance). Another intervention could focus on optimizing care and follow-up for those households with high utilization of EMS for the complaint of high blood pressure, while also educating and directing them away from future non-emergent calls. Lastly, the MOH could consider integrating data from Locator™ into the national medical database, ArMed. This would ideally prompt primary care hypertension follow-up for those patients who called an ambulance for high blood pressure, as this information would be available to their primary health care physician.

### Study Strengths and Limitations

This is the first study to evaluate and describe ambulance calls for high blood pressure in Armenia. The dataset included all ambulance calls logged into Locator™ at every state-funded ambulance station in Armenia across 6.5 years. There were some notable issues with missing data. Among 54 total ambulance stations, 14 were removed from analysis due to previously identified issues with reporting (see Arzoumanian et al. for more information about missing data in the Locator™ dataset).(13) The present study finds that data in Yerevan was unreliably logged between 2017-2019, as visualized in Figure 1.

Finally, the Locator™ software has a set list of patient complaints that can be logged in the system. Patient concerns that are not included in this list are not documented in the electronic database (notably, “headache” and “lightheadedness”). This introduces the possibility that some patients calling for high blood pressure may have reported one or more additional symptoms which were not included in our analysis; however, the majority of red flag symptoms are listed in the database (e.g. chest pain, shortness of breath).

## Conclusion

Hypertension is a major cause of premature death globally, and Armenia has one of the highest hypertension prevalence rates in the world. Nearly 29% of adults calling for an ambulance in Armenia list high blood pressure as their sole complaint, and only 9.2% of these calls result in ambulance transfer to a hospital for further evaluation and care. These findings indicate that Armenia’s high hypertension prevalence and low rate of blood pressure control contribute a significant burden on the ambulance system, and that a large proportion of ambulance resources are utilized for non-emergency cases as defined by the physician’s decision to transport the patient to a facility for definitive care.

Policymakers should prioritize these findings and consider implementing short and long term public health interventions to lessen the burden of high blood pressure calls on Armenia’s ambulance system. Future studies should investigate ambulance physician practice patterns in the treatment of patients calling EMS for high blood pressure, and investigate the hospital course for those patients who were transported. It is also important to delineate the reasons that patients are calling EMS to understand if there is a primary care access issue versus a miscomprehension of what constitutes an emergent condition. The analysis presented in this study should be repeated using the same methodology periodically to evaluate progress after the implementation of suggested interventions.

## Data Availability

No data was generated by this study. The study utilized a database generated by Locator™ software and provided by Armenia’s Ministry of Health. While the dataset is not available to the public online (as it is maintained by the government of Armenia), data can be accessed through collaboration with officials at Armenia’s Ministry of Health and Locator™ representatives. We, the authors of the study, cannot publish the dataset, as it would be an ethical breach in confidentiality between our team and Armenia’s Ministry of Health. Additionally, any population and geographic data generated in Armenia has the potential for misuse by bad actors in the ongoing geopolitical conflicts of the region.

